# The attributable fraction of respiratory syncytial virus among patients of different age with influenza-like illness and severe acute respiratory illness in a high HIV prevalence setting, South Africa, 2012-2016

**DOI:** 10.1101/2022.11.22.22282618

**Authors:** Jocelyn Moyes, Stefano Tempia, Sibongile Walaza, Meredith L. McMorrow, Adam L. Cohen, Florette Treurnicht, Orienka Hellferscee, Nicole Wolter, Anne Von Gottberg, Halima Dawood, Ebrahim Variava, Kathleen Kahn, Shabir A. Madhi, Cheryl Cohen

## Abstract

**Introduction:** The detection of respiratory syncytial virus (RSV) in upper airway samples does not necessarily infer causality of illness. Calculating the attributable fraction (AF) of RSV in clinical syndromes could refine disease burden estimates.

**Methods:** Using unconditional logistic regression models, we estimated the AF of RSV-associated influenza-like illness (ILI) and severe-acute respiratory illness (SARI) cases by comparing RSV-detection prevalence among ILI and SARI cases to those of healthy controls in South Africa, 2012-2016. The analysis, stratified by HIV serostatus, was conducted in the age categories <1, 1-4, 5-24, 25-44, 45-64, ≥65 years.

**Results:** We included 12,048 individuals: 2,687 controls, 5,449 ILI cases and 5,449 SARI cases. RSV-AFs for ILI were significant in <1, 1-4, 5-24, 25-44-year age groups: 84.9%(95% confidence interval (CI) 69.3%-92.6%), 74.6%(95%CI 53.6%-86.0%), 60.8%(95%CI 21.4%-80.5%) and 64.1%(95%CI 14.9%-84.9%), respectively. Similarly, significant RSV-AFs for SARI were 95.3%(95%CI 91.1%-97.5) and 83.4%(95%CI 70.9-90.5) in the <1 and 1-4-year age groups respectively. In HIV-infected persons, RSV was significantly associated with ILI cases versus controls in individuals aged 5-44 years.

**Conclusion:** High RSV-AFs in young children confirm RSV detection is associated severe respiratory illness in South African children, specifically infants. These estimates will assist with refining burden estimates and cost effectiveness models.

**Key point:** The attributable fraction of illness is key to accurate burden estimates, specifically in the presents of sensitive PCR testing. Burden and cost burden estimates are key to cost-effectiveness model for new prevention technologies in the pipeline for respiratory syncytial virus.

## Background

It is estimated that up to 33.1 million episodes of RSV-associated lower respiratory tract infection (LRTI) occur annually globally in children under-5 years of age, approximately 10% of which result in hospitalisation^1^. Of the nearly 120,000 estimated RSV-associated global deaths among children aged <5 years annually, 99% occur in low-and-middle income countries of Africa and Asia^2^. Data from South Africa confirm a substantial RSV burden in children, with estimated hospitalisation rates of 3,262/100,000 population in children aged <1 year^3^. HIV-exposed infants under 6 months of age are considered at increased risk of hospitalisation with RSV-associated LRTI compared to HIV-unexposed infants (incidence rate ratio (IRR) 1.4; 95% CI 1.3-1.6)^4^.

In addition, there may be substantial burden of RSV-associated LRTI in adults, with estimated incidence of hospitalisation 30/100,000 in the general South African population aged ≥ 5 years and from 106 to 390/100,000 population in individuals living with HIV^5^. HIV infection is a known risk factor for severe respiratory illness (SRI) in all age groups ^5^.

Most recent burden estimates have relied on sensitive polymerase chain reaction (PCR) assays to detect RSV in respiratory samples^6^. Describing the correlation between the detection of RSV on these sensitive assays and the presence of RSV disease is important to refine burden estimates. Accurate burden estimates will be important to advocate for the introduction of new RSV prevention technologies, such as vaccines and new generation monoclonal antibodies^7,8^. In addition, estimates of the attributable fraction (AF) of RSV (RSV-AF) in LRTI and influenza like-illness (ILI) will assist in building accurate cost-effectiveness models for the RSV interventions in the pipeline.

While there are some data on RSV-AF in children, there is very little information on the RSV-AF in HIV-infected and HIV-exposed uninfected (HEU) children^9,10^. The AF of RSV to disease in adults is also not well characterised. Prior to the introduction of interventions for RSV-associated illness, policy makers need accurate estimates of both in- and out-patient RSV disease burden. Burden estimates collected prior to introduction will assist with the assessment of vaccine impact.

We aimed to evaluate the AF of RSV-detection in individuals of all ages presenting with non-hospitalized influenza-like illness (ILI) or hospitalised severe acute respiratory illness (SARI). Further we looked specifically at the following groups: HIV-infected and HIV-uninfected children <5 years of age, HEU infants (aged <6 months); and lastly in HIV-infected and HIV-uninfected adults. These analyses were further stratified by age-groups <1 year, 1-4 years, 5-24 years, 25-44 years, 45-64 years and ≥65 years. We also aimed to estimate the proportion of ILI and SARI cases attributable to RSV infection after adjusting the observed detection rate of RSV for the RSV-AF.

## Methods

Cases were enrolled as part of a prospective, hospital-based, sentinel surveillance programme for respiratory illness from June 2012 through May 2016 at two sites in North West (Klerksdorp-Tshepong Hospital Complex (KTHC), Klerksdorp), and KwaZulu-Natal (Edendale Hospital (EDH), Pietermaritzburg) provinces of South Africa and primary health care clinics in the hospital catchment population^11^. Specific descriptions of enrollment are provided below.

### Controls and influenza-like illness (ILI) cases

Study-specific controls were defined as individuals who sought medical care at one of the sentinel clinics for non-respiratory, non-gastrointestinal symptoms and who did not have a recorded temperature or history of fever within the last 14 days. Most of the controls were selected from non-urgent outpatient service visits including dental clinics, family planning, immunization and well-baby visits, voluntary HIV counselling and testing, or acute care for minor injuries. Two controls, one living with and one without HIV, were enrolled weekly in each clinic among age groups: <1 year, 1–4 years, 5–24 years, 25–44 years, 45–64 years, and ≥ 65 years. ILI cases were enrolled at the same clinics. ILI was defined as illness in an outpatient of any age who had a recorded temperature ≥38°C or a history of fever and cough of ≤10 days duration.

### Severe acute respiratory illness (SARI) surveillance

A SARI case was defined as an illness in a hospitalised individual meeting the following age-specific criteria. A SARI case in children 3 months to <5 years of age was defined as any child with physician-diagnosed acute LRTI with symptom duration ≤10 days, including bronchopneumonia, pneumonia, bronchitis, bronchiolitis, and pleural effusion irrespective of signs and symptoms; we also included infants aged 2 days to <3 months with physician-diagnosed sepsis. A SARI case in individuals aged ≥5 years was defined as any individual with recorded temperature ≥38°C or history of fever and cough.

### Study procedures

Detailed procedures for the surveillance program have been described^11^. Briefly, dedicated surveillance staff systematically screened potentially eligible outpatients and hospitalized individuals and collected a clinical specimen and detailed history of symptoms and other medical conditions from consenting persons meeting surveillance case definitions. Individuals with ILI who were referred to hospital on the day of enrolment were excluded from the analysis.

### Determination of HIV status

HIV status was determined from one of following sources: (i) in hospital/clinic voluntary counselling and testing by rapid HIV test according to standard clinical protocols (ii) patient records or (iii) if necessary (with consent), a dried blood spot tested at the National Institute for Communicable Diseases (NICD), Johannesburg. Testing for individuals aged ≥18 months was done by an HIV ELISA, and PCR testing was conducted on samples from children aged <18 months.

### HIV-exposed-uninfected infants

HIV exposed-uninfected (HEU) infants were those aged <6 months born to a mother who was living with HIV, and in whom the HIV infection status of the child was confirmed to be negative. These infants were enrolled at the same surveillance sites as the cases and controls above.

### Samples collection and virus detection

Nasopharyngeal aspirates were collected from children aged <5 years and for individuals aged ≥5 years both oropharyngeal and nasopharyngeal swabs were collected. The specimens were placed in universal transport medium (Copan, Murruieta, California, USA) and stored at 4°–8°C at the surveillance site prior to being transported to NICD for testing. Testing was conducted within 72 hours of collection. All specimens were tested for RSV and nine other respiratory viruses (influenza virus A and B, parainfluenza types 1, 2 and 3, adenovirus, human metapneumovirus, enteroviruses and rhinovirus). Testing was done on a multiplex real-time reverse transcription polymerase chain reaction assay during 2012-2014^12^. From 2015 a combination of the commercial Fast Track Diagnostics (FTD) Flu/HRSV assay (Fast Track Diagnostics, Luxembourg) and Allplex Respiratory Assay, panel 2 and 3 (Seegene, Seoul, Korea) in 2015 were used to test for other respiratory viruses.

### Statistical analysis

We used an unconditional logistic regression to estimate the AF of RSV-associated SARI and ILI by comparing the RSV detection rate observed in our surveillance among ILI and SARI to those of controls ^13,14^. These estimates were adjusted for HIV infection and co-infections with the other respiratory viruses tested. The analysis was implemented overall (all age groups), and stratified by HIV serostatus in the following age categories: <1 year, 1-4 years, 5-24 years, 25-44 years, 45-64 years, and≥65 years. HIV infection and age group were also included as covariates in the non HIV-stratified overall model and the models implemented among children aged <5 (<1 and 1-4 years) and person aged ≥5 years (5-24, 25-44, 45-64 ≥65 years). Significant associations were considered at p<0.05.

The adjusted odds ratios obtained from these models were used to estimates the attributable fraction (AF) by applying the formula: 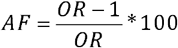. We also estimated the RSV detection rate associated with illness among individuals with ILI and SRI (*Prev*_*Illness*_) from the observed detection rate (*Prev*_*Observed*_) as follows: *Prev*_*Illness*_ = *Prev*_*Observed*_ \**AF*.

The statistical analysis was conducted using STATA version 14.1 (StataCorp, College Station, Texas, USA).

### Ethical considerations

Approval for the hospital-based surveillance was obtained from the University of the Witwatersrand Human Research Ethics Committee (HREC) for the Klerksdorp site and form the University of KwaZulu-Natal Human Biomedical Research Ethics Committee (BREC) for the Pietermaritzburg site; protocol numbers M081042 and BF157/08, respectively. The ILI and control enrolment protocol was approved by HREC and BREC (protocol numbers M120133 and BF080/12, respectively). This surveillance was deemed non-research by the US Centers for Disease Control and Prevention (non-research determination number: 2012-6197).

## Results

### Study population

We enrolled 12,048 individuals of whom 99% (12,008) had RSV test results. This included 2,687 controls, 5,449 with ILI and 3,912 with SARI.

Overall, the RSV detection positivity was 2% (52/2,687) in controls, 5% (303/5,449) in ILI cases and 14% (553/3,912) in SARI cases. The highest positivity was in children aged <1 year with SARI (31%; 413/1,328). Among ILI cases, detection was highest in children aged <1 year (13%; 78/621). RSV detection was similar among controls (3%; 12/389 and 3%; 17/552) in age groups <1 year and 1-4 years, respectively (Table 1). HIV results were available for 95% (11,445/12,048) of enrolled participants.

**Table 1:** Attributable fraction (AF) and AF-adjusted prevalence of respiratory syncytial virus (RSV) among individuals in a study of the AF of RSV detection in mild and severe illness, Klerksdorp and Edendale, South Africa, June 2012 to May 2016

| All enrolled participants |  |  |  |  |  |  |
| --- | --- | --- | --- | --- | --- | --- |
| Influenza-like Illness (ILI) |  |  |  | Severe Acute Respiratory Illness (SARI) |  |  |
|  | Observed RSV detection rate | Attributable Fraction (AF) % (95% Confidence interval (CI)) | AF adjusted prevalence % | RSV detection rate | AF, % (95% CI) | AF- adjusted prevalence % |
| <1 | 78/621 (13) | **84.9 (69.3-92.6)** | 11 | 413/1328 (31) | **95.3 (91.1-97.5)** | 30 |
| 1-4 | 116/1101 (11) | **74.6 (53.6-86.0)** | 8 | 101/786 (13) | **83.4 (70.9-90.5)** | 11 |
| 5-24 | 43/1531 (3) | **60.8 (21.4-80.5)** | 2 | 6/287 (2) | 25.6 (0-73.8) | 1 |
| 25-44 | 49/1532 (3) | **64.1 (14.9-84.9)** | 2 | 16/842 (1) | 26.1 (0-72.6) | <1 |
| 45-64 | 15/561 (3) | 69.6 (0-90.8) | 2 | 11/492 (2) | 52.6 (0-87.9) | 1 |
| 65+ | 2/103 (2) | 48.0 (0-94.7) | 1 | 6/177 (3) | 80.6 (0-96.8) | 2 |
| Individuals living with HIV |  |  |  |  |  |  |
|  | Observed RSV detection rate | ILI |  | Observed RSV detection rate | SARI |  |
|  |  | AF, % (95%CI) | AF-adjusted prevalence, % |  | AF, % (95%CI) | AF-adjusted prevalence, % |
| <1 | 0/8 (0) | 0 (0-92.5) | Not determined | 3/20 (15) | 0 (0-71.9) | Not determined |
| 1-4 | 1/19 (5) | 75.8 (0-96.5) | 4 | 9/91 (10) | 72.1 (0-95.9) | 7 |
| 5-24 | 12/230 (5) | **78.1 (36.5-92.5)** | 4 | 5/114 (3) | 55.4 (0-87.7) | 3 |
| 25-44 | 29/865 (3) | **76.6 (13.6-93.7)** | 2 | 16/722 (2) | 61.8 (0-90.0) | 1 |
| 45-64 | 6/227 (3) | 75.7 (0-96.1) | 2 | 8/299 (3) | 80.4 (0-97.6) | 2 |
| 65+ | 0/10 (0) | No RSV | NA | 3/33 (9) | 87.4 (0-99.4) | 8 |
| **HIV-uninfected individuals** |  |  |  |  |  |  |
|  |  | **ILI** |  |  | **SARI** |  |
|  | **Observed RSV**<br>**detection rate** | **AF, % (95%CI)** | **AF-adjusted**<br>**prevalence, %** | **Observed RSV detection**<br>**rate** | **AF, % (95%CI)** | **AF-adjusted prevalence, %** |
| <1 | 68/574 (12) | **87.5 (72.2-94.4)** | **11** | 371/1109 (33) | **96.7 (93.0-98.5)** | 32 |
| 1-4 | 112/1038 (11) | **77.7 (56.6-88.5)** | **9** | 91/656 (14) | **84.6 (70.3-92.0)** | 12 |
| 5-24 | 27/1158 (2) | 39.0 (0-74.1) | 1 | 1/123 (1) | 0 (0-78.7) | **Not determined** |
| 25-44 | 18/588 (3) | 30.9 (0-76.0) | 1 | 0/93 (0) | 0(0-53.5) | **Not determined** |
| 45-64 | 7/301 (2) | 60.8 (0-91.0) | 1 | 0/162 (0) | 0 (0-91.3) | **Not determined** |
| 65+ | 1/84 (1) | 46.0 (0-94.5) | 0.5 | 3/129 (2) | 64.3 (0-94.8) | 1 |

### Attributable fractions

#### Influenza-like illness

RSV-AFs in ILI cases were significantly different from zero in the following age groups: <1 year (84.9%, 95% CI 69.3%-92.6%), 1-4 years (74.6%, 95% CI 53.6%-86.0%), 5-24 years (60.8%, 95% CI 21.4%-80.5%) and 25-44 years (64.1%, 95% CI 14.9%-84.9%). In older age groups, the detection of RSV in ILI cases was not significantly attributable to disease (45-64 years, 69.6%, 95% CI 0%-90.8% and ≥65 years, 48.0%, 95% CI 0%-94.7%) (Table 1, Figure 1a-c).

**Figure 1:**
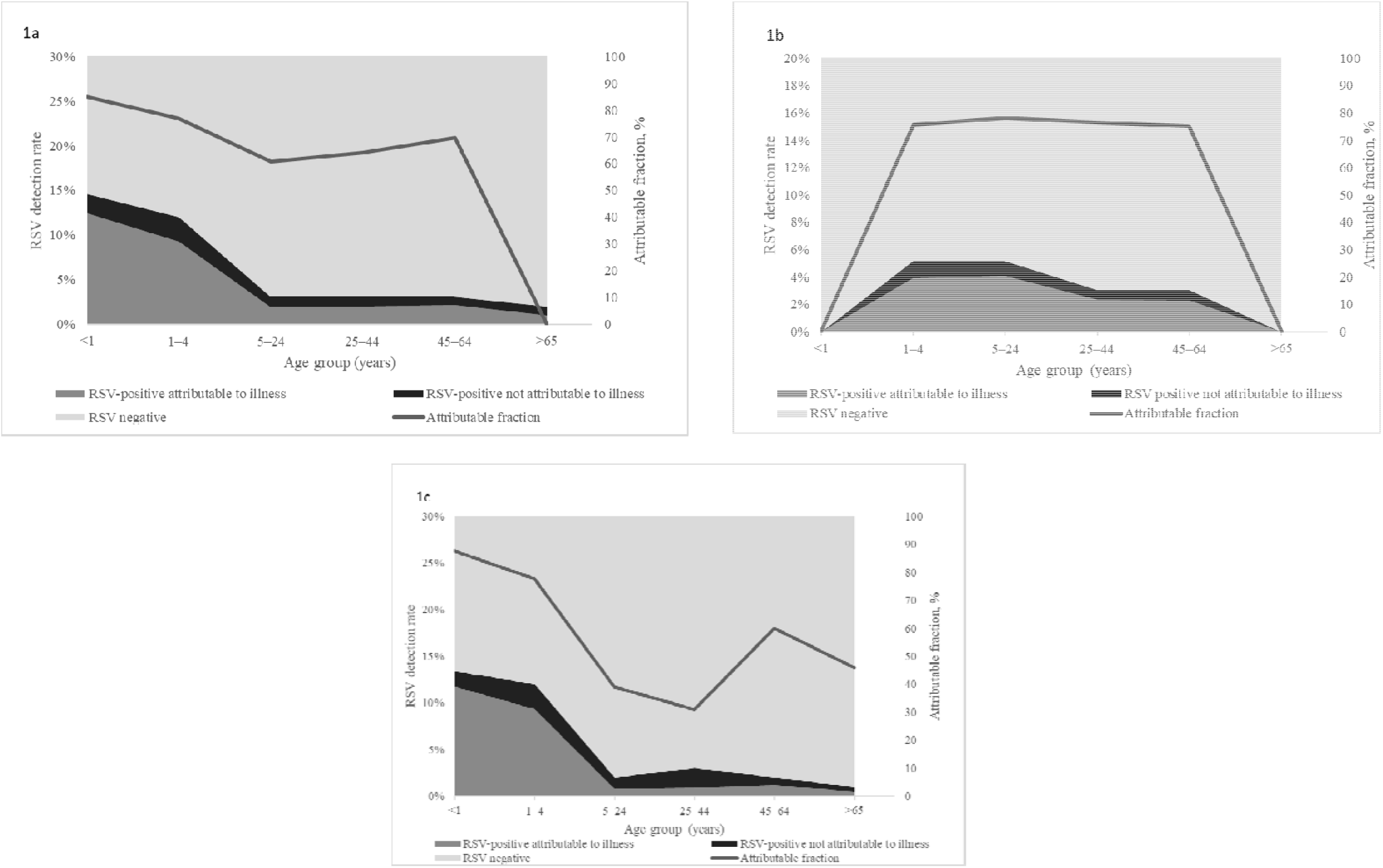
Attributable fraction of respiratory syncytial virus (RSV) and proportion of RSV-positive cases attributable to illness and RSV-positive cases not attributable to illness in individuals with influenza-like illness (ILI) (1a: Overall, 1b: HIV-infected and 1c: HIV uninfected), in two provinces South Africa 2012-2016.

#### Severe acute respiratory illness

The RSV-AFs for SARI were significantly different from zero in the age groups <1 year (95.2%, 95% CI 90.9%-97.5) and 1-4 years (82.9%, 95% CI 70.2-90.1) (Table 1, Figure 2a). This association was also seen in HIV-uninfected individuals with similar RSV-AFs observed in HIV-uninfected infants aged <1 year with SARI (96.7%, 95% CI, 93.0%-98.5%) (Table 1, Figure 2a-c). The RSV-AF was not significantly different from zero in the age group ≥65years irrespective of HIV status (80.6%, 95% CI, 0%-96.6%) and similarly in the age groups 5-24 years, 25-44 years and 45-64 years (25.6%, 95% CI 0%-73.8%; 26.1%, 95% CI 0%-72.6% and 52.6%, 95% CI, 0%-87.9%, respectively) (Table 1, Figure 2a-c)

**Figure 2:**
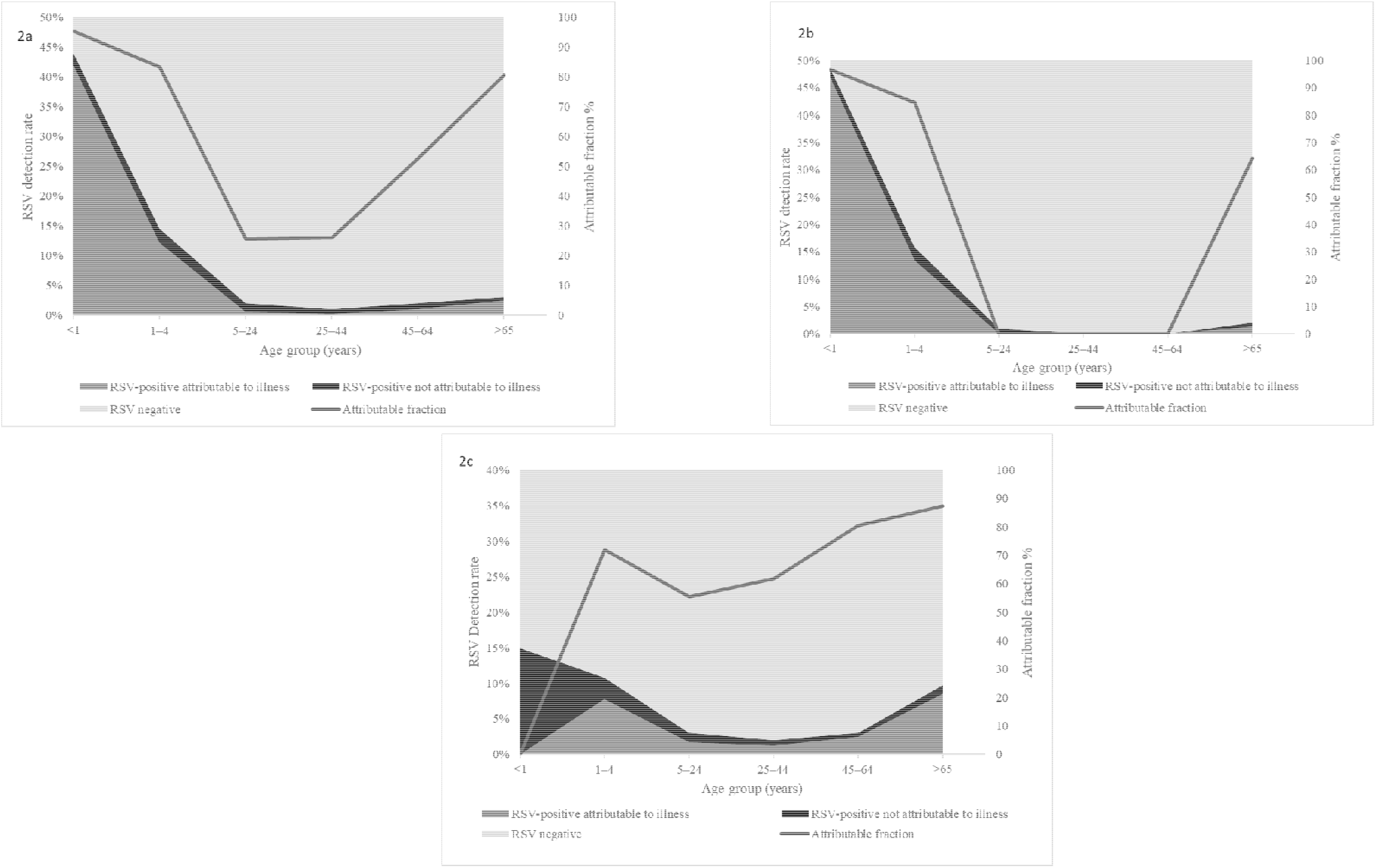
Attributable fraction of respiratory syncytial virus (RSV) and proportion of RSV-positive cases attributable to illness and RSV-positive cases not attributable to illness in individuals with severe acute respiratory illness (SARI) (2a: Overall, 2b: HIV-infected and 2c: HIV-uninfected), in two provinces South Africa 2012-2016. (Foot note RSF-AF in age group 5-24, 25-44 and 45-54 were not estimated for HIV-uninfected individuals as no cases were detected in these age groups)

#### HIV-infected individuals

Among HIV-infected individuals, the RSV-AFs for ILI were significantly different from zero in the 5-24 and 25-44-year age groups only 78.1%, 95% CI, 36.5%-92.5% and 76.6%, 95% CI 13.6%-93.7%, respectively). We were not able to calculate the RSV-AFs for ILI and SARI in the <1-year age groups due to no RSV observations occurring in HIV-infected infants with ILI and SARI (Table1, Figure 1b and 2b).

#### HIV-exposed infants

The RSV-AF, for HIV-exposed-uninfected infants (HEU) <6 months of age with SARI, was significantly greater than zero (89.9%, 95% CI 69.2%-96.7%). The RSV-AF for ILI in HEU <6 months of age was not significantly different from zero (64.2%, 95% CI 0%-90.1%). The RSV-AF for HIV-unexposed-uninfected (HUU) infants aged <6 months was significantly different from zero across the clinical syndromes (ILI 88.0%, 95% CI 54.2%-96.8% and SARI 97.0%, 95% CI 86.0%-99.3%) (Table 2).

**Table 2:**
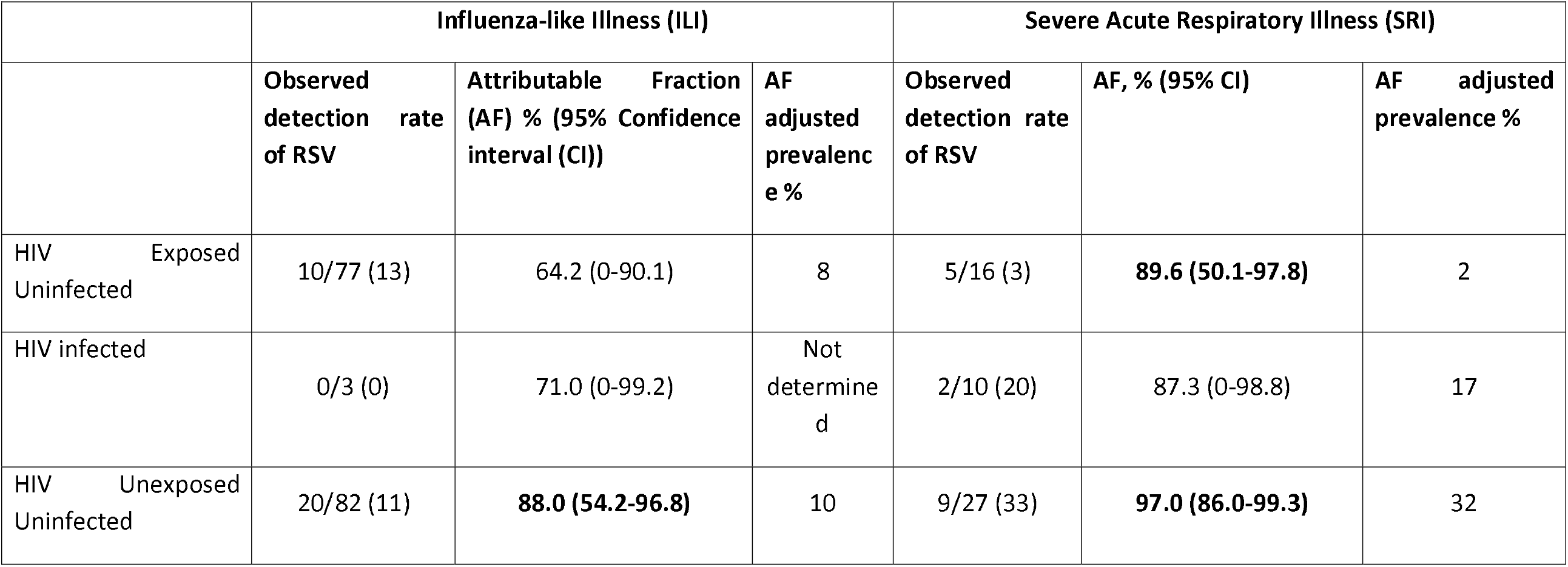
Attributable fraction (AF) and AF-adjusted prevalence of respiratory syncytial virus (RSV) among HIV exposed-uninfected (HEU), HIV-infected and HIV-unexposed uninfected (HUU) infants aged <6 months, Klerksdorp and Edendale, June 2012 to May 2016

## Discussion

We were able to describe RSV-AFs that were significantly different from zero in ILI in all age groups except for the older adult age groups (45-64 years and ≥65 years). We confirmed the high RSV-AF in children with SARI with significant RSV-AF in children aged <1 year. In addition, we documented no RSV-AFs in adults with RSV-associated SARI which were significantly different from zero. These associations were similar for adults living with HIV. In HIV-uninfected individuals aged ≥5 years, the detection of RSV was not significantly associated with illness. The RSV-AFs were lower in HEU infants aged <6 months, as compared to the HUU, but higher than HIV-infected infants aged <6 months.

In children the RSV-AF was high in all age groups, specifically in the children <1 year presenting with SARI confirming the findings in other studies^3,15^. Detection of RSV was significantly associated with illness in the children presenting with ILI aged <1 year and 1-4 years. Several studies in children confirm a strong association between the detection of RSV and disease that we describe in this analysis. Although there are many different study designs and case definitions of SARI the results are remarkable similar across studies^9,10,15,16,17,18^. An interim analysis of a subset of data presented in this study showed similar results to our analysis for those aged <5 years^14^. In the Drakenstein study, a case-control study, conducted in South Africa, of the association between the detection of respiratory viruses and pneumonia, RSV detection was associated with pneumonia (odds ratio (OR)) 8.5, 95% CI; 4.2-15.4 and very strongly associated with very severe illness OR 25.3, 95% CI 3.3-191.6^10^. A study in western Kenya described an association between RSV detection and disease in children aged <5 years with pneumonia (adjusted OR 3.0, 95% CI 1.1-8.2)^18^. The PERCH multicentre study reported that viruses account for almost two thirds of pneumonia cases (61.4%, 95% credible interval (CrI) 57.3-65.6. In addition, this study reported RSV as having the greatest aetiological fraction 31.3 95% CrI, 28.4-34.2) of all pathogens, very similar to our results^17^. Additional evidence is presented in a systematic review and meta-analysis by Shi et al where the authors assign a causal attribution OR of RSV in children of 9.8, 95% CI 4. 9-19.3 and an AF of 90%, 95% CI 85%-95%^15^.

In our assessment, among individuals living with HIV we describe that RSV detection is significantly associated with illness in individuals aged 5-24 years and 25-44 years for ILI. In HIV-infected children aged <5 years we did not see an association between the detection of RSV and illness, this is likely due to the very low prevalence of HIV in this age group, limiting our ability to detect significant associations. We did not detect any cases of RSV-associated ILI or SARI in HIV-infected individuals <1 year. HEU infants are at increased risk of LRTI and hospitalisation with RSV-associated LRTI, the lower RSV-AF in this group compared to HIV-unexposed uninfected infants suggest that other pathogens are contributing proportionately more to the disease in this group of infants^4^.

In individuals aged ≥5 years with SARI the point estimates of RSV-AF suggest that RSV may be associated with illness however in many age groups the RSV-AFs were not significantly higher in cases compared to controls. Although older age (≥65 years) is considered a risk factor for RSV-associated SARI, we do not demonstrate a significant RSV-AF in this group of individuals^19^. This may be due to the low number of elderly individuals presenting at health care facilities in our setting or because there is not truly an association in this age group.

When our RSV-AF data are displayed graphically by age group, U-shaped distributions are seen among SRI cases, with high RSV-AF in young children, dropping in younger adults and rising again in the elderly (although point estimates of AF were not significant in some groups). Similar to the shape of the distribution for the incidence of hospitalisation with RSV-associated SRI^5,20,21^. By comparison the influenza-AF curve among individuals with SARI has lower estimates for children <1 year and higher estimates for the ≥65 year age group^22^. Conversely the RSV-AF in individuals with ILI forms a more bell-like shape with highest AF in the age groups 25-64 years. Data on the causal association of viral detection in LRTI in adults are sparse. In the EPIC study in the United States of America the incidence of RSV-associated pneumonia in adults was estimated at 7/100,000 population 95% CI, 5%-9%, lower than the estimate in our setting but without AF estimation, therefore the numbers are not comparable to our study^20,21^.

## Conclusion

RSV is strongly associated with respiratory illness particularly in young South African children. While point estimates suggested an elevated AF for RSV-associated SARI among the elderly, we were not able to conclusively demonstrate an association between RSV detection and illness. More data are needed on the RSV-AF to illness among the elderly. These data are of interest to clinicians and policy makers. In addition, the calculation of the AF will improve the estimates of burden and cost-effectiveness models. These estimates of AF could be used to motivate for the introduction of vaccines and new generation monoclonal antibodies^8^.

## Data Availability

All data produced in the present study are available upon reasonable request to the authors

## Disclaimer

The findings and conclusions in this report are those of the author(s) and do not necessarily represent the official position of the funding agencies.

## Contributions

Jocelyn Moyes: Study design, implementing field work, data analysis and manuscript preparation. Stefano Tempia: Study design, statistical advise, manuscript preparation. Sibongile Walaza: Managed field work, data management and contribution to preparing the manuscript. Meredith L. McMorrow: Supported manuscript writing and data anlysis. Adam L. Cohen: Data management, analysis and writing. Florette Treurnicht: Sample testing, verifying laboratory results, contributed to manuscript writing. Orienka Hellferscee: Sample testing, verifying laboratory results, contributed to manuscript writing. Nicole Wolter: Sample testing, verifying laboratory results, contributed to manuscript writing. Anne Von Gottberg: Verifying laboratory results, contributed to manuscript writing. Halima Dawood: Assisted with field work set up, access to hospitals and writing the manuscript. Ebrahim Variava: Assisted with field work set up, access to hospitals and writing the manuscript. Kathleen Kahn: Assisted with field work set up, access to hospitals and writing the manuscript. Shabir A. Madhi: Study design, analysis and manuscript review. Cheryl Cohen: Study design, statistical support, data analysis and manuscript writing.

